# Urban rail transport and SARS-CoV-2 infections: an ecological study in Lisbon Metropolitan Area

**DOI:** 10.1101/2020.09.18.20195776

**Authors:** Milton Severo, Ana Isabel Ribeiro, Raquel Lucas, Teresa Leão, Henrique Barros

**Affiliations:** EPIUnit - Instituto de Saúde Pública, Universidade do Porto, Rua das Taipas, n° 135, 4050-600 Porto, Portugal; Departamento de Ciências da Saúde Pública e Forenses e Educação Médica, Faculdade de Medicina, Universidade do Porto, Porto, Portugal

**Keywords:** SARS-CoV-2, Transport policy, Disease transmission, coronavirus, urbanhealth

## Abstract

**Introduction:** Large number of passengers, limited space and shared surfaces can transform public transportation into a hub of epidemic spread. This study was conducted to investigate whether proximity to railway stations, a proxy for utilization, was associated with higher rates of SARS-CoV-2 infection across small-areas of Lisbon Metropolitan Area (Portugal).

**Methods:** The number of SARS-CoV-2 confirmed infections from March 2 until July 5, 2020 at parish-level was obtained from the National Epidemiological Surveillance System. We used a Geographic Information System to estimate proximity to railway stations from the six railway lines operating in the area. Then, we fitted a quasi-Poisson generalized linear regression model to estimate the relative risks (RR) and corresponding 95% Confidence Intervals (95%CI).

**Results:** Between May 2 and July 5, 2020, there were a total of 17,168 SARS-CoV-2 infections in the Lisbon Metropolitan Area, with wide disparities between parishes.

Globally, parishes near one of the railway lines (Sintra) presented significantly higher SARS-CoV-2 infection rates (RR=1.42, 95%CI 1.16, 1.75) compared to those parishes located far away from railway stations, while the opposite happened for parishes near other railway lines (Sado/Fertagus), whose infection rates were significantly lower than those observed in parishes located far away from railway stations (RR=0.66, 95%CI 0.50, 0.87). However, the associations varied according to the stage of the epidemic and according to mitigation measures in place. Regression results also revealed an increasing influence of socioeconomic deprivation on SARS-CoV-2 infections.

**Conclusions:** We found no consistent association between proximity to railway stations and SARS-CoV-2 infection rates in the most affected metropolitan area of the country, suggesting that other factors (e.g. socioeconomic deprivation) might play a more prominent role in the epidemic dynamics.

## INTRODUCTION

By the beginning of 2020, societies worldwide were experiencing an unprecedented, disruptive event – the coronavirus disease 2019 (COVID-19) pandemic caused by the severe acute respiratory syndrome coronavirus 2 (SARS-CoV-2). The first confirmed case of COVID-19 in Europe was identified on January 23, 2020, and in Portugal the first case was diagnosed on March 2, 2020 (1). According to the latest data available (from September 14, 2020), there were a total of 65,021 confirmed cases of COVID-19 and 1875 deaths in Portugal.

Urban areas have been epicentres of the COVID-19 pandemic (2). In Portugal, reported SARS-CoV-2 infections are concentrated in the two main metropolitan areas of the country: Lisbon Metropolitan Area and Porto Metropolitan Area, which account for 46% and 23% of all reported infections, respectively, while concentrating 28% and 17% of the population in the country. Many who live, work, and attend school in urban areas use public transportation (metro, bus, trains) for daily commuting. Public transportation can act as a hub for epidemic spread, since there is a high number of individuals in close proximity in closed spaces, making it difficult to keep a safe distance (3). The existence of multiple surfaces, such as seats, handrails, doors, and ticket machines, shared daily by thousands of individuals, also facilitate the transfer of SARS-CoV-2 virus (4), which is believed to persist in surfaces for several hours (5). Notably, public transport is more used by low-income individuals, who do not have the possibility to opt for other modes of transportation such as a personal car (6). Adding to this, recent studies suggest that low-income individuals are more likely to be affected by most of the known risk factors for COVID-19, such as poor housing conditions, presence of comorbidities and occupation in essential sectors, such as industry, cleaning, food supply or construction (7-9).

During the lockdown period, the role of public transportation on epidemic spread was not considered particularly relevant as most countries recorded a drastic reduction in the use of public transportation (10). In Portugal, the State of Emergency was declared on March 19, 2020 and was renewed biweekly until May 2, 2020 (11). State of Emergency legislative measures enforced the closure of international borders and the suspension of non-essential services and events. Residents could leave their homes only to shop for basic needs, to take care of vulnerable people, to walk their dogs or dispose of daily residuals, and to go to work. Travelling to work was limited to those in essential jobs, and working from home was encouraged as the norm. Consequently, between March and May 2020, we observed a remarkable decrease of 70 to 80% in the utilization of public transportation (10).

As the lockdown eased and workplaces and services reopened, the use of public transport grew again, although as of September 2020, it still remained below the pre-lockdown levels (12). Lisbon Metropolitan Area is the most populated metropolitan area of the country and concentrates the highest proportion of residents that use public transport on a daily basis, namely urban trains. According to the latest Population and Housing Census (2011), 7.6% of the population from Lisbon Metropolitan Area uses trains as their main mode of transportation versus 2.9% in the whole country (13).

Coincidentally, after the lockdown eased, from June onwards, Lisbon Metropolitan Area recorded the largest growth in the number of SARS-CoV-2 infections and in July three quarters of all daily cases were reported in residents of that area. Lay media and other stakeholders started to question whether the upsurge in SARS-CoV-2 infections in the metropolitan area could be related with the large number of passengers that use Lisbon Urban railway services on a daily basis to commute.

A study conducted in China looked at the factors influencing the number of imported cases from Wuhan and the spread speed and pattern of the pandemic, and found that the presence of an airport or high-speed railway station was associated to the speed of the infection spread, although its link with the number of confirmed cases was weak (14). Another study, also focused on Wuhan, reported a strong association between travel by train to six major Chinese cities and the number of SARS-CoV-2 cases (15). Yet, these studies did not evaluate the role of urban short-duration train trips, typically used for daily commuting, on the number of SARS-CoV-2 infections. In addition, little is known on the role of public transport on transmission following the application of widespread containment or mitigation measures.

This ecological study was conducted to investigate whether proximity to urban railway stations, a proxy for utilization in daily commuting, was associated with higher rates of SARS-CoV-2 infection across small-areas from Lisbon Metropolitan Area, between March and July 2020.

## 2. Materials and Methods

### 2.1. Study design

This ecological study used Lisbon Metropolitan Area parishes (smallest administrative territorial unit) as observation units and compared the SARS-CoV-2 infection rates between the parishes closer and parishes farther from a train station, operationally defined below, while taking into account the specific train line that runs through the closest station.

### 2.2. Study area

The Lisbon Metropolitan Area NUT III (Nomenclature of Territorial Units for Statistics III) lies on the centre-south region of Portugal and includes 18 municipalities and 118 parishes. It is the most populated metropolitan area of the country and, according to the latest population estimates (31 December 2018), holds 2.86 million inhabitants. It should be noted that, although the geographical overlap is not complete, the Lisbon Metropolitan Area holds 78% of the population from the Lisbon Health Administration Region (Administração Regional de Saúde de Lisboa e Vale do Tejo, ARSLVT), to which reported infection cases are assigned.

### 2.3. SARS-CoV-2 infections

We used data on the cumulative number of SARS-CoV-2 infections in Lisbon Metropolitan Area, according to the parish of occurrence, from March 2 until July 5, 2020. Data was obtained from National Epidemiological Surveillance System (SINAVE) and provided by Directorate-General of Health (Direção-Geral da Saúde, DGS). SINAVE is a real-time electronic platform used by public, private and social healthcare institutions in Portugal to collect data on communicable diseases and other public health risks (16). The information collected is based on international standards for disease surveillance, recommended by the European Centre for Disease and Control and by the World Health Organization, and reported on the electronic form provided by SINAVE on https://sinave.min-saude.pt (16). After the submission of the notification by an authorized user, the data is made available in real-time for the local, regional and national health authority. The data is validated consecutively, by hierarchical level, to assure the validity of the reported information and to avoid duplicate cases.

### 2.4. Train station network and geospatial procedures

The list of railway stations in the Lisbon Metropolitan Area and corresponding railway lines was obtained on the websites from CP - Comboios de Portugal (https://www.cp.pt/passageiros/pt) and Fertagus (https://www.fertagus.pt/).

When this study was conducted (7 July 2020), there were a total of six railway lines operating in the study area. Four belonged to Comboios Urbanos de Lisboa (Linha de Azambuja, Linha do Sado, Linha de Sintra and Linha de Cascais) and two lines belonged to Comboios Regionais/Suburbanos (Linha do Oeste and Linha do Sul-Fertagus), including a total of 71 railway stations depicted in Figure 1.

**Figure 1.**
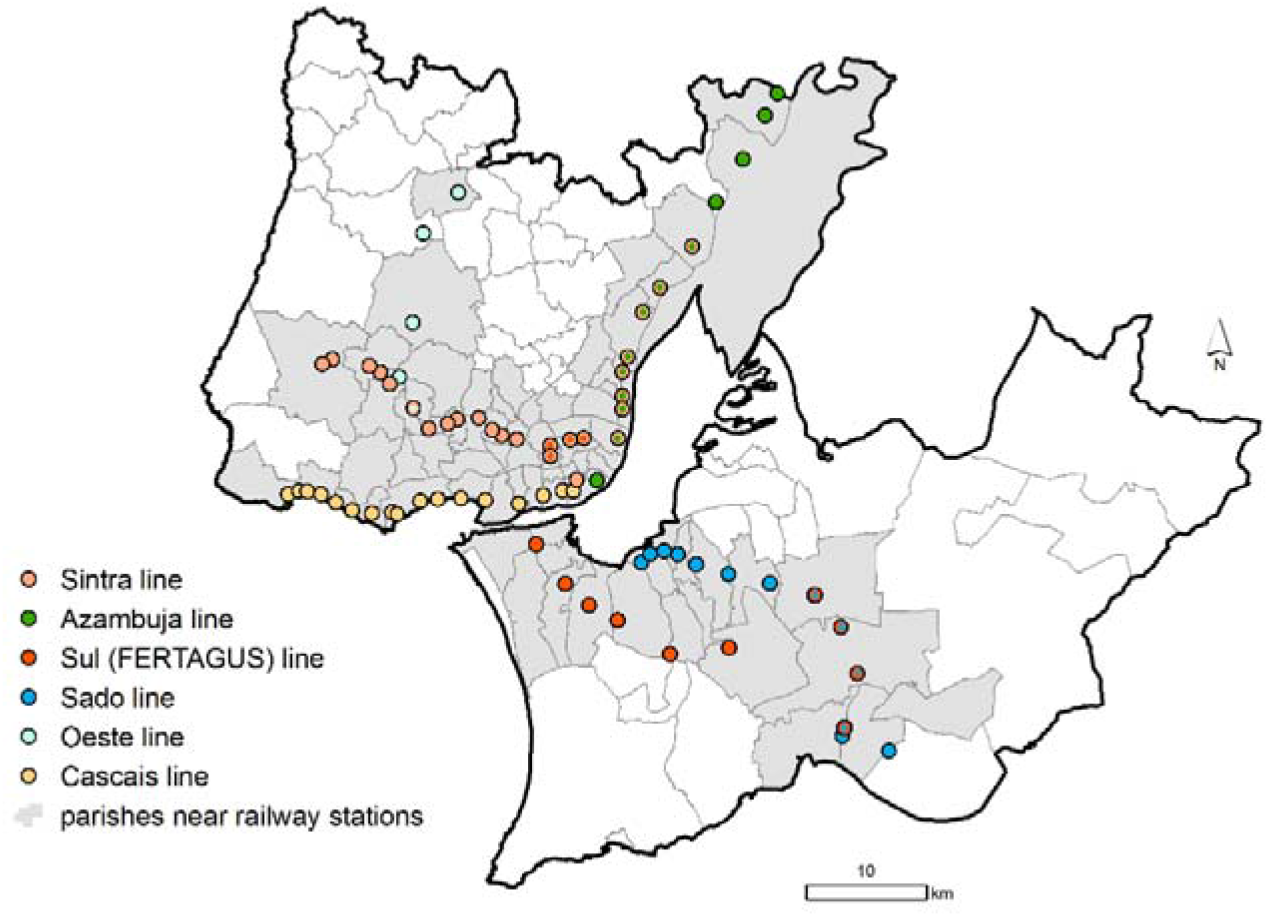
Location of the railway stations from Lisbon Metropolitan Area in 5 July 2020.

Using the official administrative cartography as a base map (Carta Administrativa Oficial de Portugal version 2019, CAOP 2019 (17)) and a Geographic Information System (ArcGIS 10.7.1), we created a 100×100 meter grid covering all the parishes belonging to Lisbon Metropolitan Area. Afterwards, using the centroid of each grid cell as origin, we computed the Euclidian distance (i.e. straight line) from that centroid to the nearest railway station. With the resultant distance matrix, we computed the shortest distance from each parish to the nearest railway station. Because each parish can be close to various stations, served by different railway lines, we assigned each parish to the line that ran through the nearest station. Then, we grouped the parishes into: parishes near railway stations (those at a mean minimum distance equal or shorter than 3000 meters, n=76) and parishes farther from railway stations (those at a mean minimum distance larger than 3000 meters, n=42). The parishes classified as near railway stations are depicted in gray color in Figure 1.

### 2.5. Statistical analysis

To estimate the magnitude of the association between SARS-CoV-2 infection rates and proximity to railway station at the parish-level (closer vs. farther), we fitted a quasi-Poisson generalized linear regression model with the log of the population as offset to estimate the relative risks (RR) and corresponding 95% Confidence Intervals (95%CI). To control for confounding, models were adjusted for socioeconomic deprivation assessed by the European Deprivation Index (EDI). The EDI was constructed in three steps, fully described elsewhere (18), and resulted from the weighted sum of the following standardized variables at parish-level: percentage of non-owned households; households without indoor flushing; households with five rooms or less; individuals with blue-collar (manual) occupations; individuals with low education level (≤6th grade); non-employers; unemployed looking for a job; and foreign residents.

The analysis was stratified according to four periods each representing a stage in the epidemic spread and mitigation measures: March-July (the whole analysis period 02/03 to 05/07), March (the early stage of the epidemic and transition to State of Emergency, 02/03 to 31/03), April (State of Emergency, 1/04 to 30/04), May (end of the State of Emergency and beginning of State of Alert, 01/05 to 31/05) and June-July (State of Alert, 01/06 to 05/07). Statistical analysis was performed using R software version 4.0.0.

## 3. RESULTS

Between March 2 and July 5, 2020 there was a total of 17,168 SARS-CoV-2 infections in the Lisbon Metropolitan Area: 1272 in March, 3257 in April, 4566 in May and 8073 in June/July. Infection rates across the Lisbon Metropolitan Area varied substantially ranging from 26 cases per 100,000 inhabitants in Santo Isidoro (Mafra) to 1725 cases per 100,000 in Alvalade (Lisbon), in the whole period.

Table 1 presents the relative risks comparing infection rates in the parishes near railway stations, according to railway line, with those from parishes located far away.

**Table 1.** Associations (Relative Risk, RR, and 95% Confidence Intervals, 95%CI) between railway station proximity and COVID-19 infection rates.

|  | March-July | March | April | May | June-July |
| --- | --- | --- | --- | --- | --- |
| Parishes far away from railway stations | Ref (1.00) | Ref (1.00) | Ref (1.00) | Ref (1.00) | Ref (1.00) |
| Parishes near railway stations from Azambuja line | 1.15 (0.90, 1.49) | 1.26 (0.77, 2.04) | 1.11 (0.74, 1.65) | 1.19 (0.86, 1.65) | 1.06 (0.80, 1.40) |
| Parishes near railway stations from Cascais line | 0.99 (0.75, 1.29) | **1.60 (1.03, 2.51)** | 1.14 (0.76, 1.69) | 0.77 (0.52, 1.13) | 0.89 (0.65, 1.20) |
| Parishes near railway stations from Oeste line | 1.12 (0.68, 1.75) | 0.80 (0.24, 2.01) | 0.88 (0.36, 1.83) | 1.16 (0.60, 2.06) | 1.18 (0.69, 1.89) |
| Parishes near railway stations from Sado/Fertagus line | **0.66 (0.50, 0.87)** | 0.72 (0.43, 1.18) | **0.65 (0.42, 0.98)** | 0.71 (0.50, 1.01) | **0.61 (0.45, 0.82)** |
| Parishes near railway stations from Sintra line | **1.42 (1.16, 1.75)** | 1.41 (0.95, 2.12) | **1.44 (1.05, 2.01)** | 1.27 (0.96, 1.68) | **1.41 (1.13, 1.78)** |
| Socioeconomic deprivation (European Deprivation Index) | **1.08 (1.05, 1.11)** | 0.97 (0.92, 1.01) | **1.05 (1.01, 1.09)** | **1.10 (1.07, 1.14)** | **1.10 (1.07, 1.12)** |

Significant associations are depicted in bold letters.

Globally, parishes near the Sintra railway line presented significantly higher SARS-CoV-2 infection rates (RR 1.42 95%CI 1.16, 1.75) compared to those parishes located far away from railway stations, while the opposite happened for parishes near the Sado/Fertagus lines, whose infection rates were significantly lower than those observed in parishes located farther away from railway stations (RR 0.66 95%CI 0.50 0.87). However, this pattern changed through the course of the epidemic and according to the measures in place at each time.

In the earlier stage of the epidemic, during March, parishes located near the Cascais railway line presented significantly higher infection rates than those located farther away (RR 1.60 95%CI 1.03, 2.51). In April, when the entire country was under a lockdown, compared to parishes farther from railway stations, parishes near railway stations from Sado/Fertagus line presented significantly lower infection rates (RR 0.65 95%CI 0.42, 0.98), while parishes located near the Sintra line presented significantly higher infection rates (RR 1.44 95%CI 1.05, 2.01). During May, when lockdown measures started to ease, no significant associations between railway station proximity and SARS-CoV-2 infection rates were observed. During June/July, as in April, parishes near railway stations from Sado/Fertagus line presented significantly lower infection rates (RR 0.61 95%CI 0.45, 0.82), while parishes located near Sintra line presented significantly higher infection rates (RR 1.41 95%CI 1.13, 1.78).

Regression results also revealed an increasing influence of socioeconomic deprivation on SARS-CoV-2 infection rates. By March, the socioeconomic deprivation index was not significantly associated with parishes’ infection rates, while in April and in May, we observed a positive association between socioeconomic deprivation and SARS-CoV-2 infection rates (April: RR 1.05, 95%CI 1.01, 1.09; May: 1.10 95%CI 1.07, 1.14).

## 4. DISCUSSION

Our results showed an inconsistent association between railway station proximity and SARS-CoV-2 infection rates, as both negative and positive associations were observed depending on the railway line and time period. We found that the most at risk areas changed throughout the epidemic and we observed a time-dependent effect of socioeconomic deprivation on area-level SARS-CoV-2 infection rates.

During April, June/July and globally throughout the COVID-19 epidemic period, we observed that parishes near railway stations from the Sintra line had significantly higher SARS-CoV-2 infection rates than parishes far away from railway stations. The Sintra line is one of the most used lines from the Lisbon Metropolitan Area guaranteeing the daily commuting of the population from Sintra and Amadora (the latter is the most densely populated municipality of the country) to the Lisbon municipality (the capital). The Sintra line is currently the busiest in the country (and one of the busiest suburban lines in Europe), carrying several tens of thousands of passengers every day at around 14 trains per hour. During the COVID-19 epidemic, physical distancing was strongly encouraged by health authorities, meaning people should keep about 2 meters or more apart from others. However, trains, such as those from the Sintra line, tend to work at maximum capacity during peak periods, making it difficult to implement such physical distancing. However, simultaneously, parishes near railway stations from Sado/Fertagus lines presented significantly lower SARS-CoV-2 infection rates than those far away from train stations. Sado/Fertagus lines are very busy as well, since they connect the southern parishes of the Lisbon Metropolitan Area located on the left side of the Tagus river estuary to the capital, and carry several tens of thousands of passengers on a daily basis.

Despite this overall pattern, at the earlier stages of the epidemic, the most affected parishes were those crossed by the Cascais line. At this stage, most reported cases were imported and were associated with national individuals coming back from international events (e.g. Milan fashion fairs), Carnival holidays and from snow resorts, many of them in Northern Italy. The Cascais municipality, despite being socioeconomic heterogeneous, is one of the wealthiest municipalities in Portugal (19) and an important tourist destination (20), which explains why it constituted an high-rate area at the earlier stages of the epidemic.

Our findings seem to exclude a direct and consistent association between proximity to railway stations and SARS-CoV-2 infections. However, the lack of association we observed between variables on an aggregate level do not necessarily represent risk at an individual level. Therefore, we cannot exclude the possibility that some individuals may have acquired the infection inside the train or at railway stations. Hence, reasonable distancing between passengers should be ensured, the use of face mask should be promoted, and disinfection of surfaces (seats, handrails, doors, and ticket machines) should be strengthened to inactivate the virus (21).

Though this was not the main objective of the study, we also observed a consistent increase in the influence of socioeconomic deprivation throughout the epidemic period and after adjusting for vicinity of railway stations. At the initial stage of the epidemic, no differences in infection rates according to socioeconomic deprivation were observed while, from April onwards, we observed roughly 10% increase in SARS-CoV-2 infection rates per unit increase in the socioeconomic deprivation index. Socioeconomic and ethnic inequalities in COVID-19 distribution and mortality have been reported in the UK (22), Brazil (23) and in the USA (24). Economically disadvantaged people are more likely to live in overcrowded houses, a risk factor for respiratory infections (25); to have unstable working conditions and incomes, being more affected by the economic recession caused by the epidemic (25); to have comorbid conditions, which may hamper the immune system’s ability to combat the infection (25) and to have lower access to healthcare (26). Additionally, disadvantaged individuals tend to be employed in occupations that do not provide opportunities to work from home during lockdowns (25). This is a very plausible set of explanations for the increase in the socioeconomic inequalities in SARS-CoV-2 infection rates that we observed from April onwards.

This study has limitations that need to be considered. First, the data and analyses were derived from an ecological approach due to the lack of information of individual use of rail transport, weakening causal inference at the individual level. Ecological designs also limit our capacity to control for confounding, meaning that other factors (for which information was unavailable) rather than proximity to railway stations or socioeconomic deprivation may partially explain our findings (e.g. other modes of transportation, types of activities taking place in each area, population ethnic composition, etc.). Second, our study may be affected by the Modifiable Areal Unit Problem (MAUP) (27), which happens when the number of spatial units (the scale) used to define the same area affects the study conclusions. If the geographical units are large, it is more likely that associations found at the aggregate level will diverge from the same associations found at individual level leading to the so-called ecological fallacy (28). In our study, we used the smallest geographical unit available to minimize this problem. A third issue, is the Uncertain Geographic Context Problem (UGCoP) (29). Case data is available according to the parish of occurrence, but focusing only on occurrence location may introduce uncertainty in research results, because people may spend a considerable amount of time in other parishes and may acquire the disease in these locations (e.g. work, transportation, etc.) (30). Finally, case data only includes cases reported to the national surveillance system, which may not be enough to fully comprehend the true magnitude of the COVID-19 pandemic. Although the true number of undetected cases is still to be ascertained, in Europe, the ratio of the total estimated cases to the observed cases was found to around 2.3 (31).

In conclusion, we found no consistent association between proximity to railway stations and SARS-CoV-2 infection rates in the most affected metropolitan area of the country, suggesting other factors, namely neighbourhood socioeconomic deprivation, play a more prominent role in the epidemic dynamics. Nevertheless, our findings do not imply that safety measures in public transportation can be relaxed – proper surface disinfection, physical distancing, and mass mask use should keep being promoted. To guide measures of epidemic control, individual-level studies, namely through the adoption of case-control designs, are recommended to better understand in which locations (e.g. work, school, shopping) there is a higher risk of acquiring the infection.

## Data Availability

Aggregated data can be made available upon request to the corresponding author.

## ACKNOWLEDGMENTS

The authors are grateful to Direção-Geral da Saúde for providing the data from the National Epidemiological Surveillance System (SINAVE).

